# Identification of High Likelihood of Dementia in Population-Based Surveys using Unsupervised Clustering: a Longitudinal Analysis

**DOI:** 10.1101/2023.02.17.23286078

**Authors:** Amin Gharbi-Meliani, François Husson, Henri Vandendriessche, Eleonore Bayen, Kristine Yaffe, Anne-Catherine Bachoud-Lévi, Laurent Cleret de Langavant

## Abstract

**Background:** Dementia is defined by cognitive decline that affects functional status. Longitudinal ageing surveys often lack a clinical diagnosis of dementia though measure cognitive and function over time. We used unsupervised machine learning and longitudinal data to identify transition to probable dementia.

**Methods:** Multiple Factor Analysis was applied to longitudinal function and cognitive data of 15,278 baseline participants (aged 50 years and more) from the Survey of Health, Ageing, and Retirement in Europe (SHARE) (waves 1, 2 and 4–7, between 2004 and 2017). Hierarchical Clustering on Principal Components discriminated three clusters at each wave. We estimated probable or “Likely Dementia” prevalence by sex and age, and assessed whether dementia risk factors increased the risk of being assigned probable dementia status using multistate models. Next, we compared the “Likely Dementia” cluster with self-reported dementia status and replicated our findings in the English Longitudinal Study of Ageing (ELSA) cohort (waves 1–9, between 2002 and 2019, 7,840 participants at baseline).

**Findings:** Our algorithm identified a higher number of probable dementia cases compared with self-reported cases and showed good discriminative power across all waves (AUC ranged from 0.754 [0.722–0.787] to 0.830 [0.800–0.861]). “Likely Dementia” status was more prevalent in older people, displayed a 2:1 female/male ratio and was associated with nine factors that increased risk of transition to dementia: low education, hearing loss, hypertension, drinking, smoking, depression, social isolation, physical inactivity, diabetes, and obesity. Results were replicated in ELSA cohort with good accuracy.

**Interpretation:** Machine learning clustering can be used to study dementia determinants and outcomes in longitudinal population ageing surveys in which dementia clinical diagnosis is lacking.

**Funding:** French Institute for Public Health Research (IReSP), French National Institute for Health and Medical Research (Inserm), NeurATRIS Grant (ANR-11-INBS-0011), and Front-Cog University Research School (ANR-17-EUR-0017).

## INTRODUCTION

Major neurocognitive disorder (MND), commonly known as dementia, is a clinical syndrome characterised by a decline in cognitive performance that compromises patient’s independence^1^. Repeated clinical visits and assessments reveal the progression from a healthy state to dementia. International diagnostic criteria are available to identify dementia cases. Yet, more than half of the cases in high income countries (HIC)^2^ and up to 90% in low and middle income countries (LMIC)^3^ remain undetected. For such, new methods are needed to identify dementia cases and to study dementia determinants at the level of countries or continents.

Several population-based surveys modelled on the United-States Health and Retirement Study (HRS) are conducted in multiple countries to study the impact of the transition between late-life work and retirement in aging people^4^. The “HRS family” studies offer the opportunity to compare ageing outcomes internationally^5^. Yet, in these and in many other surveys, clinical dementia status is either not available or only self-reported by participants or their families, which underestimates the number of cases.

In the absence of clinical diagnosis in population ageing surveys, unsupervised machine learning, generally used to discover clusters or patterns within datasets^6^, can identify probable dementia cases. In a previous work, we applied an unsupervised clustering method to cross-sectional data from HRS and Survey of Health, Ageing and Retirement in Europe (SHARE) to identify high likelihood of dementia^7^ based on variables related to demographics, comorbidities, functional status, mobility, cognition, and neuropsychiatric symptoms. However, applying this clustering method to cross-sectional data did not allow us to investigate longitudinal transition from normal to impaired functional status, or to assess risk factors associated with transition to dementia status.

Herein, we built a clustering analysis for identifying transition to high likelihood of dementia in population ageing surveys using repeated measurements of cognition and functional status with a modified unsupervised machine learning algorithm. Our objectives were to demonstrate that this method can identify probable dementia in population aging surveys where dementia is either poorly or non-diagnosed, and that this method is also efficient to study dementia risk factors. Three analyses were used to ascertain the internal validity of “Likely Dementia” status: (1) we compared “Likely Dementia” identification with self-reported dementia, (2) we studied the prevalence of “Likely Dementia” status according to sex and age, (3) we tested whether traditional dementia risk factors were associated with a higher risk of transition to “Likely Dementia” cluster. To demonstrate replicability, we conducted our study using SHARE survey and replicated it in the English Longitudinal Study of Ageing (ELSA).

## METHODS

### Study design and participants

We used the harmonised dataset provided by the Gateway to Global Aging^5^ of SHARE, a longitudinal panel study across multiple countries in Europe and Israel^8^. This population survey takes place every two years and follows a representative sample of individuals aged 50 years or older from each participating country. The harmonised version of SHARE consists of seven waves so far (the third being retrospective) conducted between 2004 and 2017. We included subjects from countries who have participated in SHARE since the first wave (ie, Austria, Belgium, Denmark, France, Germany, Greece, Israel, Italy, The Netherlands, Spain, Sweden, and Switzerland), aged 50 years or older with consecutive follow-ups.

### Selected variables

We used variables related to cognition and function to remain close to the DSM-5 criteria of MND. The selected variables are listed in the supplementary tables (Supplementary Table 1 & 2). All variables with more than 30% missing values were discarded and the remaining data were imputed using the imputeMFA command of the missMDA R package^9^.

### Clustering

We ran Multiple Factor Analysis (MFA) followed by Hierarchical Clustering on Principal Components (HCPC) using FactoMineR R package^10^ and longitudinal data from all waves at the same time. MFA is a principal component method that balances for differences in the number of active variables per domain by forming active groups (procedure details are shown in supplementary data). For the clustering, we retained only active groups that represented participants’ function or cognition (see supplementary tables). Each participant, at each wave, was assigned to one of the three possible clusters (ie each participant could transition from one cluster to another, from one wave to another longitudinally). The number of clusters was set at three based on previous work for identification of high likelihood of dementia^7^. At the first wave, we singled out a cluster with a high probability of dementia (named “Likely dementia” cluster) based on the impaired cognition and function detected in its participants. Any participant classified in “Likely dementia” cluster was permanently assigned to it (ie, making any incident case a prevalent one).

We took into account the attrition due to study dropout and death across waves. We applied Inverse Probability Weighting (IPW) using the ipw R package^11^. For each wave, a logistic regression model was built based on the participants’ age, sex, and country of origin characteristics collected at the previous wave. Weights were derived by inverting the product of the predicted probabilities computed by the model. We integrated those weights into both imputation and clustering methods.

### Self-reported diagnosis of dementia

We evaluated the discrimination power of our clustering method counting on its identification of “Likely dementia” status compared with the self-reported dementia status, data collected from the second wave of SHARE, using Sensitivity, Specificity and Area Under the Curve (AUC) metrics.

### Effect of age, sex, and risk factors for dementia

We computed the prevalence of “Likely dementia” status of each wave by sex and by age. Participants were divided into six age groups (under 65 years, 65–69 years, 70–74 years, 75–79 years, 80–85 years, and more than 85 years).

We examined the role of several modifiable risk factors^12^ in transitioning to “Likely dementia” cluster: low education, hearing loss, hypertension, excessive alcohol drinking, current smoking, depression, social isolation, physical inactivity, diabetes, obesity, and air pollution. Past history of traumatic brain injury was not available in the database and could not be tested. All risk factors were measured at baseline and were imputed if necessary.

We dichotomised all ordinal risk factors variables. Education level was categorised as high (upper secondary and vocational training or tertiary education) or low (less than upper secondary). For hearing loss, we used self-reported hearing capacity as a proxy considering it either being normal (excellent, very good, and good) or bad (fair or poor). Moderate and vigorous physical activity were merged into physically active (frequency: more than once per week, once per week, one to three times a month) or inactive (hardly ever or never). The remaining risk factors were treated as dichotomous as they were in the database: hypertension (ever had high blood pressure *vs* never had high blood pressure), drinking (21 units or more of alcohol per week *vs* less than 21 units of alcohol per week), smoking (current smoker *vs* non-current smoker), depression (Centre for Epidemiologic Studies Depression [CES-D] scale score greater than or equal to five *vs* CES-D scale score less than five), social isolation (participating in social activities weekly *vs* non-participating in social activities weekly), diabetes (ever had diabetes *vs* never had diabetes), obesity (Body Mass Index [BMI] ≥ 30 kg/m2 *vs* BMI < 30 kg/m2), air pollution (living in urban area *vs* living in rural area).

### Multistate models

In each wave, a participant could be classified in one of the three clusters (Cluster 1, Cluster 2 or Cluster 3; see above). Data being interval-censored, we applied multistate models using MSM package^13^ to study the impact of dementia risk factors on the risk of transition to “Likely Dementia” cluster.

We used age as the time-scale. It was calculated as the difference between birth date and interview date in years. In the multistate models, age was divided by 10 to facilitate the computational process without altering the Hazard Ratios (HR) results. Sex was considered as binary (male or female). All transitions were adjusted for sex, and transition towards “Likely Dementia” cluster was further adjusted for age. All covariates were set at baseline. For each risk factor, we computed its corresponding HR.

We checked the robustness of the multistate models in two steps. First, we considered death as a competing risk and added it as an absorbing state in the models. This was investigated in SHARE where vital status was reported consistently. Second, we excluded early prevalent and incident dementia cases by excluding participants categorised with a likelihood of dementia at first and second waves and ran multistate analyses again.

### Replication cohort

In order to confirm our results, we chose the harmonised version of ELSA^14^ as a replication cohort. It is a representative longitudinal panel study of people aged 50 years and over in England. It comprises nine waves ranging from 2002 to 2019.

### Ethical approval and guidelines

All participants provided informed consent and both studies obtained ethical approvals from local research committees. We followed both STROBE (STrengthening the Reporting of OBservational studies in Epidemiology) and MELODEM (The MEthods in LOngitudinal research on DEMentia) guidelines^15,16^ for the reporting of this study.

## RESULTS

### Identification of probable dementia

Of the initial sample of SHARE (n=30,419), we restricted our analyses to participants aged 50 years and over at baseline (n=29,102), who had consecutive follow-ups (n=15,278) (Figure 1). After running the clustering, the distribution between the clusters was uneven. At baseline, the first cluster (n=11,369) and the second (n=1,294) encompassed the majority of the sample, leaving a small part for the third cluster (n=535) (Table 1). Participants of the first and second clusters had similar baseline characteristics evoking healthy ageing. Participants of the third cluster were older (mean age 76.5 years [SD 11]), often female (n=368 [68.6%]), had lower education level (n=426 [79.6%] attained less than upper secondary education), more mobility impairment (mean mobility impairment score 4.9 [SD 1.5]), more functional impairment (mean Activities of Daily Living [ADL] score 3.1 [SD 1.7] and mean Instrumental Activities of Daily Living [IADL] score 4.2 [SD 1.8]), and more impaired cognition (mean immediate word recall test 2.6 [SD 1.9] and mean verbal fluency 10.4 [SD 6]) than participants of the first and second clusters at baseline. These characteristics corroborated that the third cluster was the one reflecting a high likelihood of dementia, thus we named it “Likely Dementia” cluster. Conversely, the first and second clusters were composed of participants considered dementia-free.

**Table 1:** Baseline characteristics of the SHARE study participants according to the three clusters identified by the algorithm. (*) Values were imputed using MissMDA package.

|  | SHARE |  |  |
| --- | --- | --- | --- |
|  | Cluster 1<br>(n=11,369) | Cluster 2<br>(n=3,374) | Cluster 3<br>(n=535) |
| Age, Years | 64.6 (9.6) | 65.2 (9.7) | 76.5 (11) |
| Sex |  |  |  |
| Female | 6,251 (55%) | 1,704 (50.5%) | 368 (68.8%) |
| Male | 5,118 (45%) | 1,670 (49.5%) | 167 (31.2%) |
| Education |  |  |  |
| Less than upper secondary education | 5,632 (49.5%) | 1,834 (54.4%) | 426 (79.6%) |
| Upper secondary and vocational training | 3,446 (30.3%) | 928 (27.5%) | 69 (12.9%) |
| Tertiary education | 2,291 (20.2%) | 612 (18.1%) | 40 (7.5%) |
| Mobility impairment score [0–7] | 1 (1.4) | 1 (1.4) | 4.9 (1.5) |
| Autonomy |  |  |  |
| ADL score* [0–6] | 0.1 (0.3) | 0.1 (0.4) | 3.1 (1.7) |
| IADL score* [0–7] | 0.2 (0.5) | 0.2 (0.6) | 4.2 (1.8) |
| Cognition |  |  |  |
| Immediate Word Recall [0–10]* | 5 (1.7) | 4.7 (1.9) | 2.6 (1.9) |
| Verbal Fluency [0–67]* | 19.7 (7.1) | 18.7 (7.3) | 10.4 (6) |

**Figure 1:**
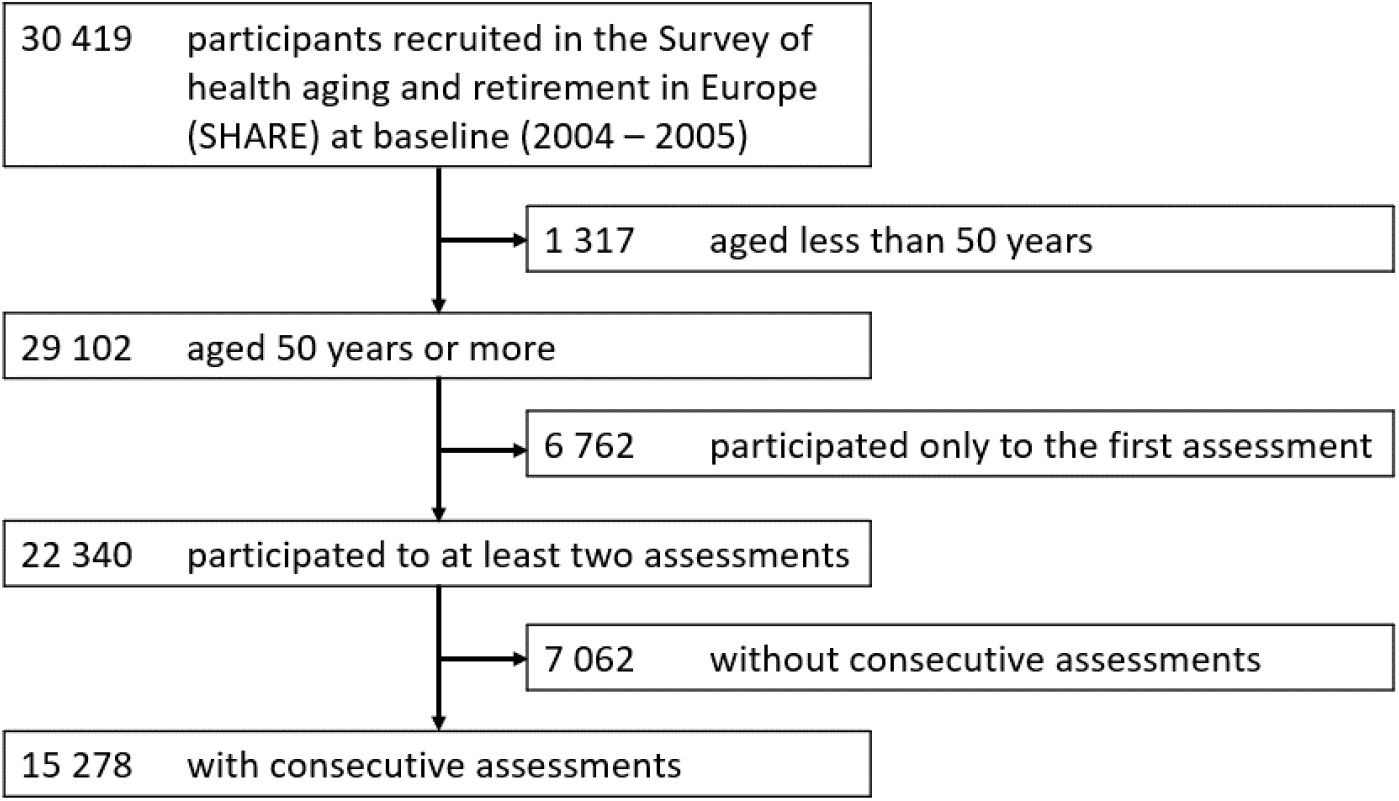
SHARE flow chart.

### Discrimination power

We compared our algorithm identification with the self-reported dementia diagnosis in the SHARE dataset, which was available from wave 2 (Table 2). Our clustering algorithm allowed the identification of a higher number of “Likely Dementia” cases compared with self-reported dementia cases. The AUC metric ranged from 0.754 (0.722– 0.787) to 0.830 (0.800–0.861), suggesting good discrimination power. Sensitivity peaked at wave 4 reaching 0.714 (0.659–0.770) then slowly decreased after. Specificity remained high (> 0.9) in all waves. Results by country are given in supplementary data (Supplementary Table 1).

**Table 2:** Comparison of self-reported dementia cases and Cluster 3 “Likely Dementia” cases; Abbreviations: AUC, Area Under the Curve; CI, confidence interval; NA, not available

| SHARE |  |  |  |  |  |  |  |  |  |  |
| --- | --- | --- | --- | --- | --- | --- | --- | --- | --- | --- |
| Wave | Number of participants | Clusters |  |  | Self-reported dementia |  |  | Metrics |  |  |
|  |  | Cluster 1 | Cluster 2 | Cluster 3 (Likely Dementia) | Missing | No | Yes | AUC (95% CI) | Sensitivity (95% CI) | Specificity (95% CI) |
| Wave 1 (2004–05) | 15,278 | 11,369 (74.4%) | 3,374 (22.1%) | 535 (3.5%) | NA | NA | NA | NA | NA | NA |
| Wave 2 (2006–07) | 15,278 | 11,433 (74.8%) | 2,832 (18.5%) | 1,013 (6.6%) | 40 (0.3%) | 14,960 (97.9%) | 278 (1.8%) | 0.805 (0.776–0.835) | 0.665 (0.610–0.721) | 0.945 (0.942–0.949) |
| Wave 4 (2010–11) | 10,008 | 7,911 (79%) | 1,406 (14%) | 691 (7%) | 21 (0.2%) | 9,735 (97.3%) | 252 (2.5%) | 0.825 (0.794–0.855) | 0.702 (0.646–0.759) | 0.947 (0.943–0.952) |
| Wave 5 (2012–13) | 8,418 | 6,429 (76.4%) | 1,312 (15.6%) | 677 (8%) | 10 (0.1%) | 8,129 (96.6%) | 279 (3.3%) | 0.794 (0.763–0.825) | 0.649 (0.593–0.705) | 0.939 (0.934–0.944) |
| Wave 6 (2014–15) | 6,485 | 4,913 (75.8%) | 987 (15.2%) | 585 (9%) | 8 (0.1%) | 6,204 (95.7%) | 273 (4.2%) | 0.755 (0.723–0.787) | 0.579 (0.520–0.637) | 0.931 (0.925–0.938) |
| Wave 7 (2016–17) | 5,533 | 3,991 (72.1%) | 1,028 (18.6%) | 514 (9.3%) | 5 (0.1%) | 5,252 (94.9%) | 276 (5%) | 0.745 (0.712–0.777) | 0.558 (0.499–0.617) | 0.931 (0.925–0.938) |

### Effect of age and sex

Older age and being female were both associated with an increased risk of entering “Likely Dementia” cluster. The prevalence of “Likely Dementia” was higher in women with approximatively a 2:1 female to male ratio across all waves (Fig 2.A). The number of “Likely Dementia” cases increased with age (Fig 2.B). For instance, at wave 2, the prevalence of “Likely Dementia” cases gradually rose with age: 1.8% in those under 65 years, 3.1% in 65–69 years, 5.9% in 70–74 years, 10.2% in 75–79 years, 18.9% in 80–85 years, and 37.4% in more than 85 years old participants.

**Figure 2:**
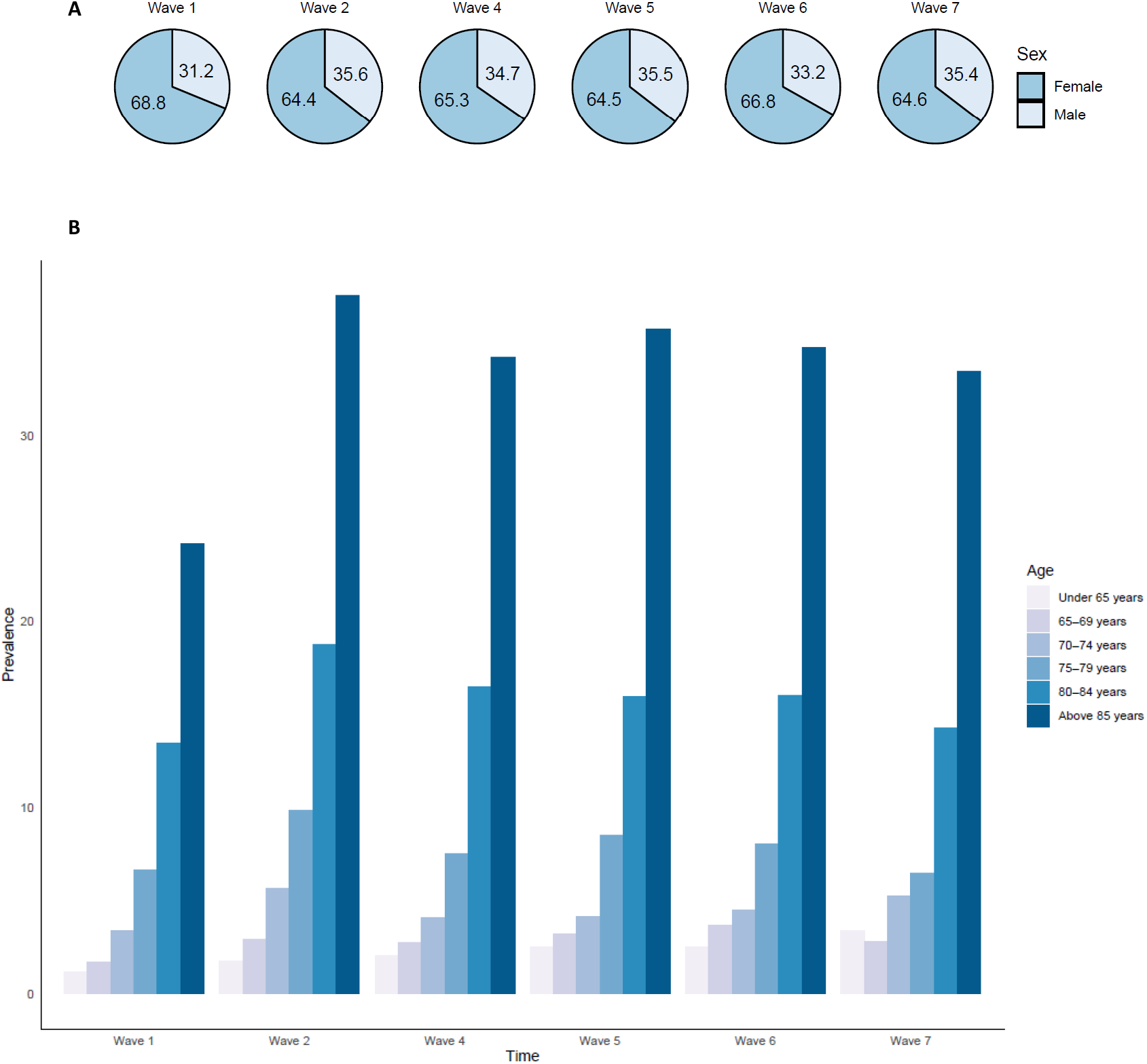
Prevalence of participants of the ‘‘Likely dementia’’ cluster by sex (2.A), and by age (2.B)

### Multistate models

To assess the associations of dementia risk factors with the risk of transitioning to ‘‘Likely Dementia’’ cluster (Table 3), we computed a multistate model (Figure 3.A). Nine of the eleven dementia risk factors chosen a priori were associated with an increased risk of transition from cluster 1 to “Likely Dementia” cluster: low education level (Hazard Ratio [HR] 1. 92% CI [1.58−2.33]), poor hearing (1.74 [1.45−2.09]), hypertension (1.35 [1.14−1.16]), smoking (1.45 [1.13−1.87]), depression (2.51 [1.06−3.07]), social isolation (1.66 [1.39−1.98]), physical inactivity (3.66 [2.97−4.51]), diabetes (2.4 [1.94−2.96]), and obesity (1.7 [1.39−2.07]). Some of these associations were also significant for transition from cluster 2 to “Likely Dementia” cluster: depression (2.39 [1.62−3.53]), social isolation (2.31 [1.51−3.53]), physical inactivity (3.21 [2.12−4.87]), and obesity (1.58 [1.08−2.32]).

**Table 3:** Multistate models for the transition to cluster 3 (“Likely dementia”) Analyses using age as time-scale. All transitions were adjusted for sex. Transition towards the third cluster (“Likely dementia”) was further adjusted for age and each risk factor individually. All risk factors were taken at baseline. Main analysis was based on a multistate model (Model 1). Sensitivity analyses were based on a multistate survival model with death as an absorbing state. First, 105 participants were removed because of inconsistencies of dates (Model 2). Second, cases identified either at the first or the second wave were removed (Model 3). Abbreviations: HR, hazard ratio; CI, confidence interval.

| SHARE |  |  |  |  |  |  |
| --- | --- | --- | --- | --- | --- | --- |
|  | Main analysis |  | Sensitivity analyses |  |  |  |
|  | Model 1 (n=15,278) |  | Model 2 (n=15,173) |  | Model 3 (n=14,278) |  |
|  | HR (95% CI)<br>(1 → 3) | HR (95% CI)<br>(2 → 3) | HR (95% CI)<br>(1 → 3) | HR (95% CI)<br>(2 → 3) | HR (95% CI)<br>(1 → 3) | HR (95% CI)<br>(2 → 3) |
| Education | **1.92 (1.58-2.33)** | 1.32 (0.91-1.9) | **1.86 (1.6-2.17)** | 1.18 (0.86-1.61) | **1.77 (1.52-2.07)** | 0.97 (0.67-1.4) |
| Hearing | **1.74 (1.45-2.09)** | 1.23 (0.85-1.79) | **1.38 (1.2-1.59)** | 1.03 (0.75-1.42) | **1.2 (1.03-1.4)** | 0.88 (0.59-1.32) |
| Hypertension | **1.35 (1.14-1.16)** | 1.24 (0.9-1.72) | **1.36 (1.2-1.55)** | 1.14 (0.86-1.5) | **1.34 (1.17-1.53)** | 1.09 (0.76-1.55) |
| Drinking (> 21 units) | 0.79 (0.55-1.14) | 0.42 (0.16-1.09) | 1.25 (0.99-1.58) | 0.36 (0.13-1.04) | **1.37 (1.09-1.73)** | 0.54 (0.21-1.41) |
| Smoking | **1.45 (1.13-1.87)** | 1.29 (0.73-2.29) | **1.64 (1.36-1.99)** | **2.23 (1.57-3.16)** | **1.7 (1.39-2.07)** | **2.68 (1.79-4.03)** |
| Depression | **2.51 (1.06-3.07)** | **2.39 (1.62-3.53)** | **2.05 (1.76-2.4)** | **1.98 (1.42-2.77)** | **1.78 (1.51-2.11)** | **1.65 (1.07-2.54)** |
| Social isolation | **1.66 (1.39-1.98)** | **2.31 (1.51-3.53)** | **1.61 (1.4-1.86)** | **1.6 (1.15-2.24)** | **1.56 (1.35-1.81)** | 1.15 (0.79-1.68) |
| Physical inactivity | **3.66 (2.97-4.51)** | **3.21 (2.12-4.87)** | **2.48 (2.07-2.97)** | **2.89 (2-4.17)** | **2.09 (1.72-2.54)** | 1.33 (0.68-2.6) |
| Diabetes | **2.4 (1.94-2.96)** | 1.32 (0.79-2.21) | **2.15 (1.82-2.54)** | 1.32 (0.85-2.05) | **2.22 (1.88-2.62)** | 0.88 (0.45-1.73) |
| Obesity | **1.7 (1.39-2.07)** | **1.58 (1.08-2.32)** | **1.65 (1.41-1.93)** | **1.43 (1.01-2.01)** | **1.76 (1.5-2.06)** | 1.32 (0.83-2.1) |
| Air Pollution | 0.84 (0.7-1.02) | 1.26 (0.83-1.9) | 0.92 (0.79-1.07) | 1.32 (0.92-1.89) | 0.94 (0.81-1.1) | 1.32 (0.83-2.08) |

**Figure 3:**
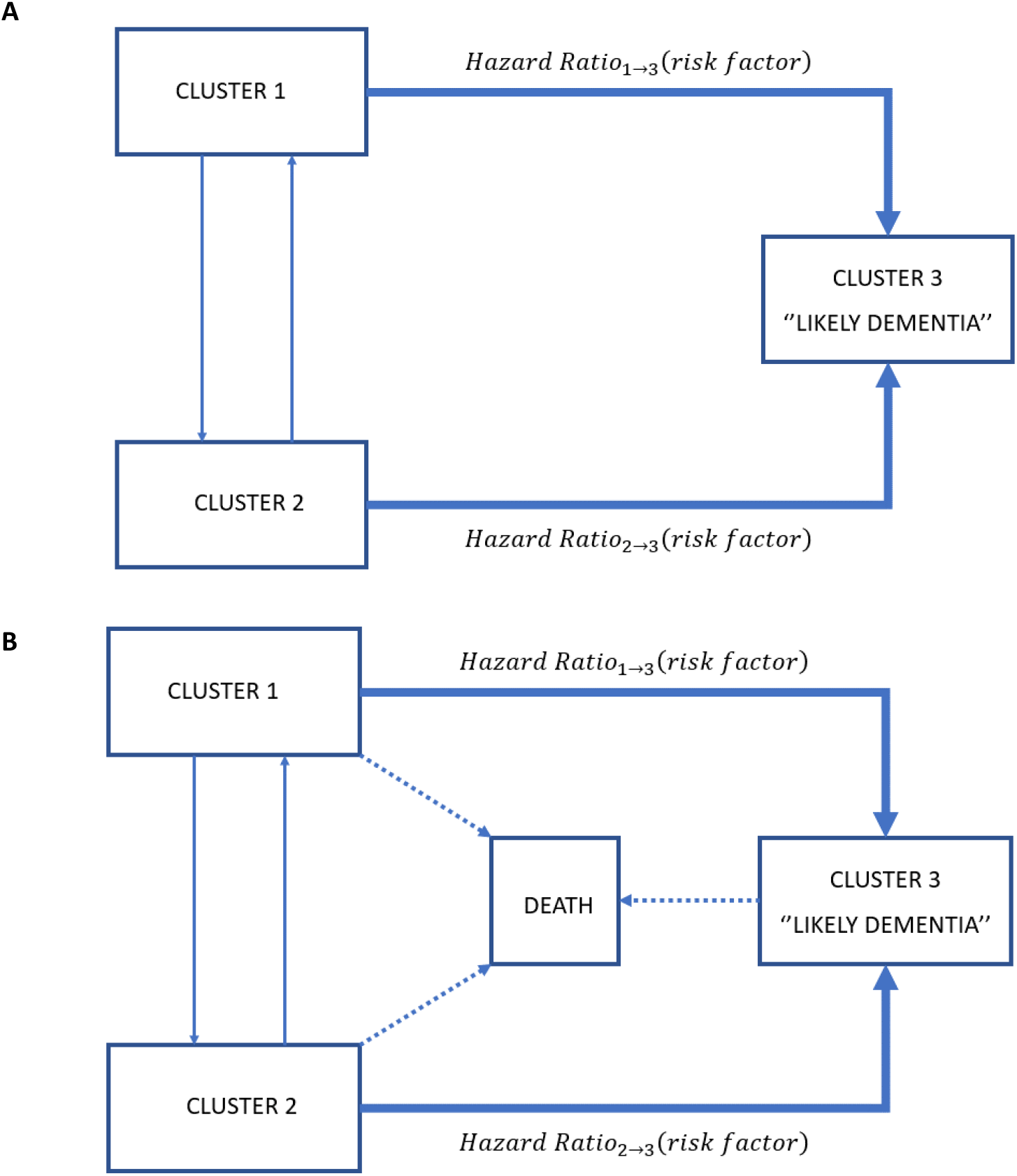
Three-state model (A) Multistate model (B) Multistate survival model.

In the first sensitivity analysis which took into account death (Figure 3.B), we excluded 105 participants due to inconsistencies between interview and death dates. All of the above-described associations between dementia risk factors and transition to “Likely dementia” cluster remained significant albeit with lower HR, except for hypertension. Of more, smoking became significantly associated with the risk of transition from cluster 2 to “Likely Dementia” cluster (2.23 [1.57−3.16]). In the second sensitivity analysis, where prevalent and incident cases at wave 1 (2004−05) and wave 2 (2006−07) (n=1,000) were further removed, HRs of transition from cluster 1 to ‘‘Likely Dementia’’ cluster did not change, but excessive alcohol drinking became a significant risk factor (1.34 [1.17−1.53]). As for transitions from cluster 2 to ‘‘Likely Dementia’’ cluster, only smoking (2.68 [1.79−4.03]) and depression (1.65 [1.07−2.54]) remained significant.

### Replication in ELSA

Of the initial sample of ELSA (n=12,099), we restricted our analyses to participants over 50 years at baseline (n=11,522) and further restricted to participants who had consecutive follow-ups (n=7,840) (Supplementary Figure 2). Overall, results obtained with ELSA participants were similar to those found in the SHARE cohort (Supplementary Table 4).

At baseline, participants of the third cluster (n=659) were more likely older (mean age 69.8 [SD 11]), more likely female (n=401 [60.8%]), of lower education level (n=423 [64.2%] attained less than upper secondary education), had more mobility impairments (mean mobility impairment score 4.9 [SD 1.5]), more functional impairment (mean ADL score 2.7 [SD 1.5] and mean IADL score 2.5 [SD 1.4]) and worse cognition (mean immediate word recall test 4.6 [SD 1.9] and mean verbal fluency 16.5 [SD 6]) than the other clusters.

Our clustering algorithm identified a higher number of “Likely Dementia” cases compared with self-reported dementia cases. Except for wave 1 (2002−03) in which the number of self-reported dementia cases was small (n=24), the algorithm identification AUC metric values were similar to those found with SHARE (Supplementary Table 5). Sensitivity and specificity were balanced.

Women were more likely to be in the “Likely Dementia” group, and prevalence of “Likely Dementia” status rose with age (Supplementary Figure 3).

Ten dementia risk factors were tested (not air pollution due to missing urbanicity data). Their associations with transition to “Likely Dementia” cluster remained similar to those found with the SHARE dataset (Supplementary Table 6) except for excessive alcohol drinking which was protective for the transition from cluster 1 to “Likely Dementia” cluster (HR 0.6 [0.43−0.83]). Four risk factors were associated with an increased risk of transition from cluster 2 to “Likely Dementia” cluster: hypertension (1.64 [1.13−2.38]), depression (2 [1.26−3.17]), physical inactivity (2.69 [1.73−4.18]), and diabetes (2.23 [1.26−3.95]). We did not take death into account in the multistate models as vital status data were not available for each wave in sensitivity analysis.

Removing prevalent and incident cases at wave 1 (2002−03) and wave 2 (2004−05) in sensitivity analysis led to similar results with few exceptions. Excessive alcohol drinking was no longer significant for the transition from cluster 1 to ‘‘Likely Dementia’’ cluster (0.79 [0.58−1.08]). Only physical inactivity remained significant for the risk of transition from cluster 2 to ‘‘Likely Dementia’’ cluster (2.02 [1.1−3.69]).

## DISCUSSION

Unsupervised clustering applied to two longitudinal population-based surveys of ageing (SHARE and ELSA) identified participants with high likelihood of dementia using longitudinal data related to functional and cognitive measures. In both surveys, this method had a good discrimination performance when compared with self-reported diagnosis of dementia. “Likely Dementia” status was more common in older participants and in women with a 2:1 sex ratio. Low education, hearing loss, hypertension, smoking, depression, social isolation, physical inactivity, diabetes, and obesity were associated with a higher risk of subsequent transition to “Likely dementia” cluster. Results for excessive alcohol drinking and air pollution were inconclusive. Applying clustering to longitudinal cohorts for the identification of high likelihood of dementia paves the way for researchers to conduct future secondary analyses on population ageing surveys worldwide.

Although supervised machine learning algorithms have already been used in population surveys to identify persons with dementia^17^, they have their limitations, eg, they require a subsample of data to be labelled “diagnosis of dementia”, and their external validity remains variable. Conversely, unsupervised machine learning may overcome such limitations as suggested in a previous cross-sectional study^7^. Here, we used an improved clustering method combining longitudinal data and a limited number of variables related to participants’ cognition and daily functions. Our clustering algorithm identified a greater number of people with a high likelihood of dementia in both SHARE and ELSA compared with self-reported dementia cases. Identifying a higher number of probable dementia cases in population ageing surveys might give a better statistical power to future studies of dementia determinants and outcomes. Moreover, this clustering method relies on cognitive and functional status data, largely available in HRS family studies and in several population ageing surveys, which makes it very suitable to apply to other ageing surveys including those in LMIC. Noteworthy, our study took into account many biases inherent to longitudinal studies, in particular attrition^18^ due to loss to follow-up or death. Internal validity was assessed using different approaches: comparison with self-reported diagnosis of dementia, impact of age and sex on dementia prevalence, and impact of known dementia risk factor on the risk of being classified as a “Likely Dementia” case. Results were obtained using data of 12 countries participating in SHARE, and then replicated in ELSA.

Nonetheless, our results should be carefully examined. Our algorithm detects a “Likely Dementia” status which cannot, by any stretch, be taken as a diagnosis of the disease without clinical validation. Future studies that compare our identification method with the recently developed cognitive assessment in HRS family cohorts using the Harmonized Cognitive Assessment Protocol (HCAP)^19^ are warranted. Our method cannot distinguish the aetiology of dementia, whether Alzheimer’s disease (AD) or other origins. Contrary to the results of our prior cross-sectional study, Cluster 1 and Cluster 2 participants were similar in terms of daily function, cognition, and mobility, yet they differed in their risk of transition to Cluster 3. However, we cannot rule out the possibility that the non-significant HRs observed for the transition from Cluster 2 to “Likely Dementia” cluster resulted from a lack of statistical power. Although this three-cluster partition remains consistent with our earlier work^7^, future investigation will test the interest of further simplification by merging the first two clusters together. The lack of biological or imaging biomarkers in this study could be seen as a limitation. Yet, biomarkers are often costly, human expert-dependent and rarely available in large population ageing studies. As for genetics, Apolipoprotein E (*APOE*)^20^ and polygenic scores^21^ are associated with a higher risk of AD, but the role of genetic factors in explaining future risk of dementia remains modest^21,22^. Although most dementia risk factors were associated with a higher risk of being assigned a “Likely Dementia” status, results for excessive alcohol drinking were ambiguous. We observed a deleterious drinking effect in SHARE, whereas it was protective in ELSA. Excessive drinking has been entangled for the brain damage it causes^23^, yet its exact relationship with dementia risk is debated since alcohol thresholds and time of exposure differ between studies^24,25^. The association between air pollution and dementia was inconclusive in SHARE and could not be explored in ELSA. Urbanicity (ie, geographical variation between urban and rural areas) was used as a proxy for air pollution as proposed recently^12^. Yet, people living in rural areas have shown higher rates of dementia compared with their urban counterparts ^26,27^. Switching to quantifiable pollution markers (fine particulate matter or ozone) that have been linked to an increased risk of dementia^28^ is more than desirable.

Unsupervised clustering is an efficient method to detect people with probable dementia in population ageing surveys using their cognitive and functional characteristics in a longitudinal setting. This approach opens new perspectives for the analyses of population data sets already available worldwide in HIC and LMIC to better compare and understand dementia determinants and outcomes.

## Data Availability

ELSA raw data can be downloaded via the UK Data Service (https://ukdataservice.ac.uk/find-data/). SHARE raw data can be accessed on their website (http://www.share-project.org/data-access.html).

http://www.share-project.org/data-access.html

## Contributors

AGM and LCL contributed to the conceptualisation and design of the study. AGM analysed the data. FH granted statistical methods expertise. HV helped with the computational analyses. AGM and LCL wrote the first draft. All authors reviewed and approved the final version of the manuscript. AGM decided to submit this manuscript for publication.

## Data sharing

All the data we used are publicly available. We used the harmonised versions of SHARE and ELSA provided by the Gateway to Global Aging Data (https://g2aging.org/). ELSA raw data can be downloaded via the UK Data Service (https://ukdataservice.ac.uk/find-data/). SHARE raw data can be accessed on their website (http://www.share-project.org/data-access.html).

## Acknowledgements

The Gateway to Global Aging Data project is developed by Centre for Economic and Social Research (CESR) at University of Southern California (https://cesr.usc.edu/). The project is funded by National Institute on Aging, National Institutes of Health (R01 AG030153, RC2 AG036619, R03 AG043052, R24 AG048024)

SHARE was funded by the European Commission, through FP5 (QLK6-CT-2001-00360), FP6 (SHARE-I3: RII-CT-2006-062193, COMPARE: CIT5-CT-2005-028857, SHARELIFE: CIT4-CT-2006-028812), FP7 (SHARE-PREP: GA 211909, SHARE-LEAP: GA 227822, SHARE M4: GA 261982, DASISH: GA 283646), and Horizon 2020 (SHARE-DEV3: GA 676536, SHARECOHESION: GA 870628, SERISS: GA 654221, SSHOC: GA 823782), and by DG Employment, Social Affairs and Inclusion (VS 2015/0195, VS 2016/0135, VS 2018/0285, VS 2019/0332 and VS 2020/0313). Additional funding from the German Ministry of Education and Research, the Max Planck Society for the Advancement of Science, the US National Institute on Aging (U01_AG09740-13S2, P01_AG005842, P01_AG08291, P30_AG12815, R21_AG025169, Y1-AG-4553-01, IAG_BSR06-11, OGHA_04-064, HHSN271201300071C, and RAG052527A) and from various national funding sources is gratefully acknowledged.

ELSA was funded by the National Institute on Aging (R01AG017644) and by a consortium of UK government departments: Department for Health and Social Care; Department for Transport; Department for Work and Pensions, which is coordinated by the National Institute for Health Research (NIHR, 198-1074). Funding has also been provided by the Economic and Social Research Council (ESRC).

First and foremost, we would like to express our gratitude to Julie Josse for all the methodological recommendations she gave for this project.

We would like to thank both Bioinformatics services of *Institut Mondor de Recherche Biomédicale* (IMRB) and *Ecole Normale Supérieure* (ENS) for their help in employing computer clusters for our analysis.

## Supplementary documents

Multiple Factor Analysis (MFA) is a principal component method similar to Principal Component Analysis (PCA) but, compared to PCA, it balances the influence of groups of variables. In our case, it balances the influence of two components, participants’ function and cognition, which have equal importance in the clinical definition of dementia (data structure for SHARE is given in Supplementary Figure 1). Then, as PCA, it gives a representation of individuals in such a way that individuals are close on the representation if they have close values from the point of view of all the variables of all the groups. Subsequently, clustering can be performed on the MFA results. Full description of the variables used for SHARE and ELSA studies are available in Supplementary Table 1 and 2, respectively.

**Supplementary Figure 1:**
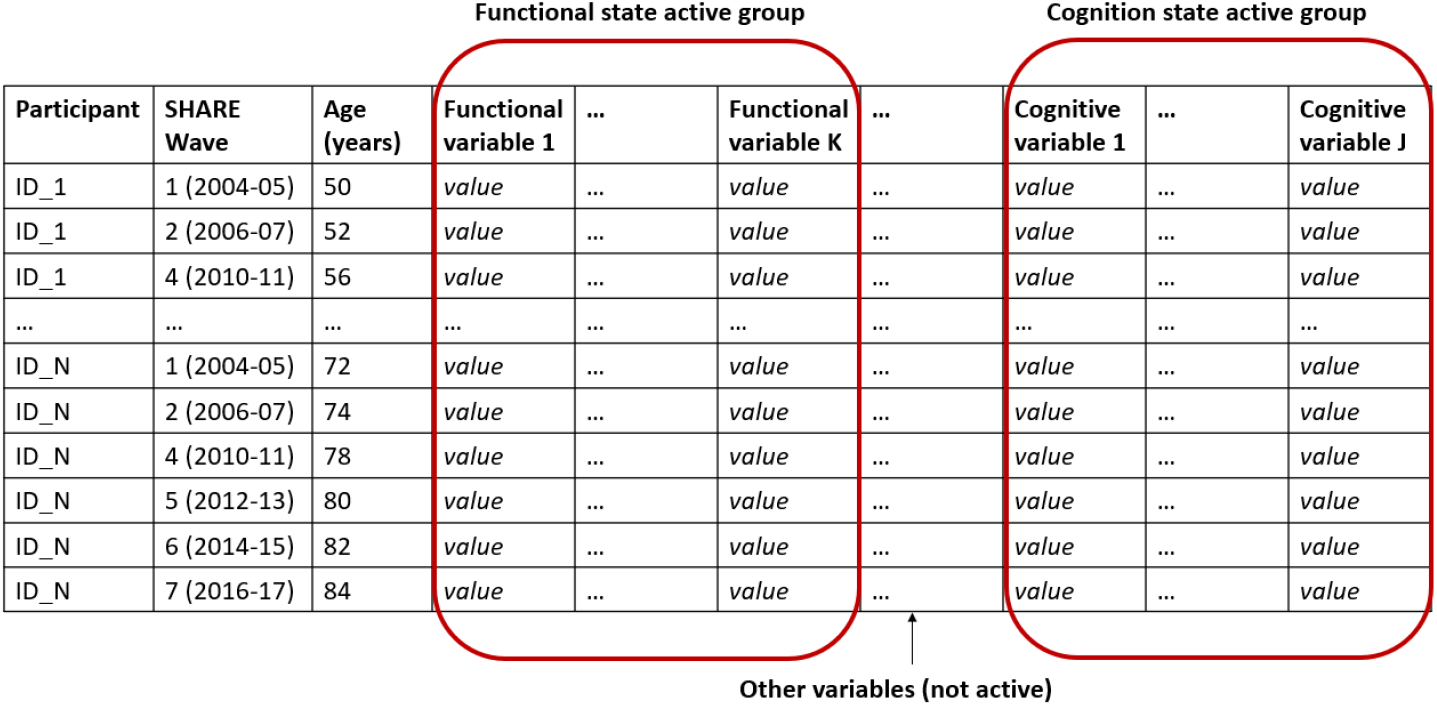
Data structure in SHARE. Each participant (from 1 to N) was seen multiple times (at least twice). Variables informative of functional status (from 1 to K) and variables informative of cognition (from 1 to J) formed the two active groups in the Multiple Factor Analysis (MFA).

**Supplementary Table 1:**
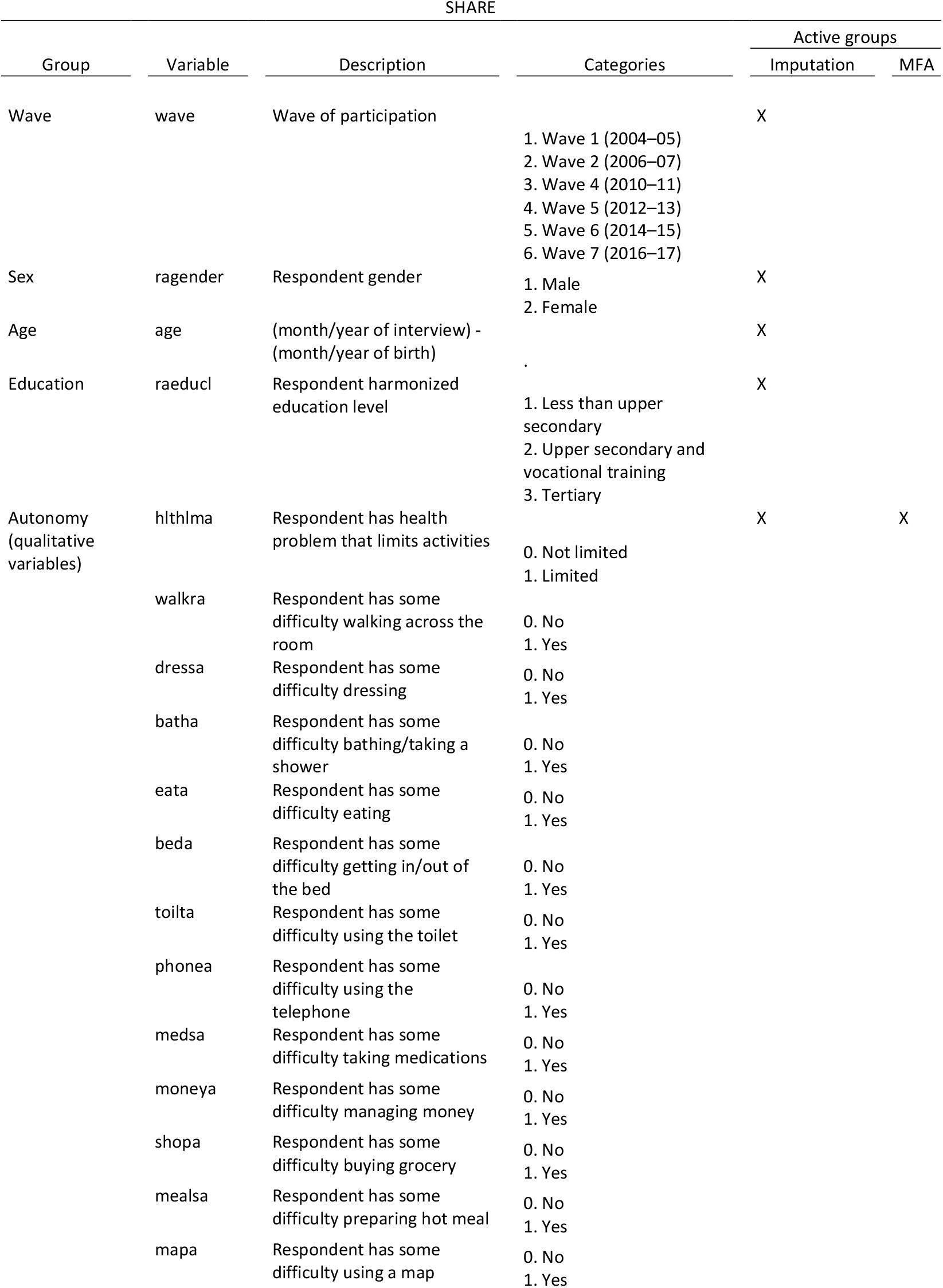
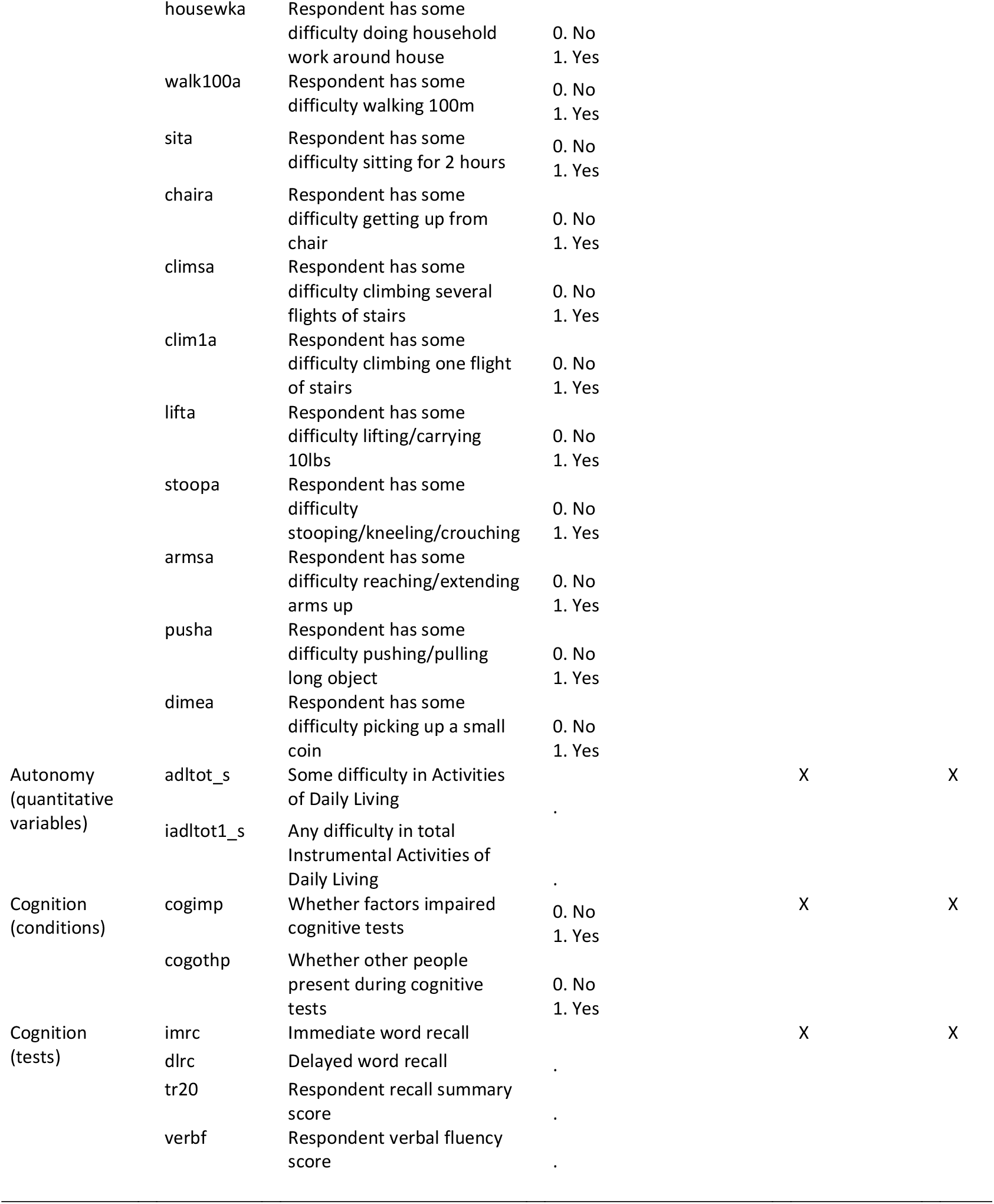
Summary of variables used for both imputation and Multiple Factor analysis (MFA) in SHARE.

**Supplementary Table 2:**
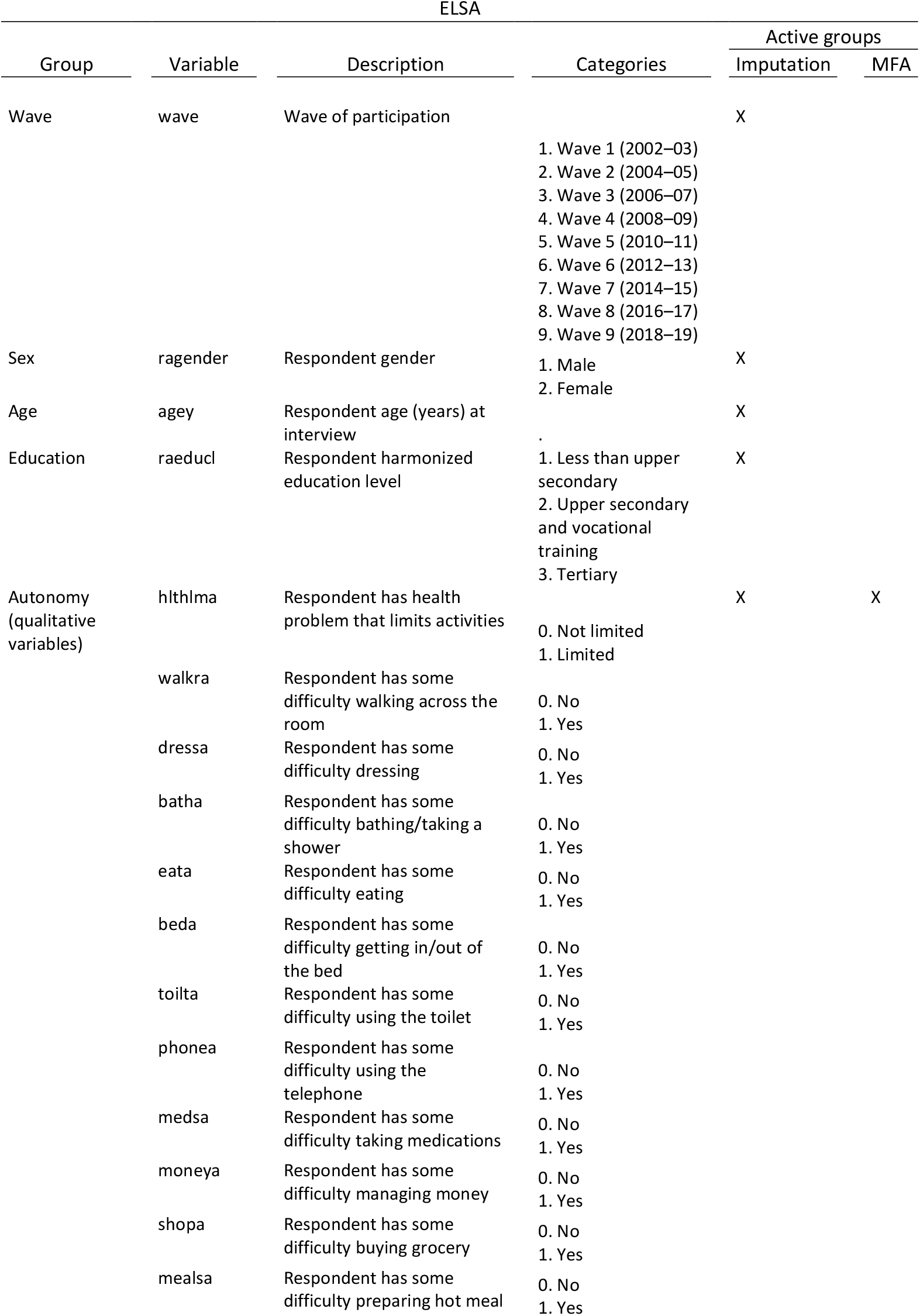
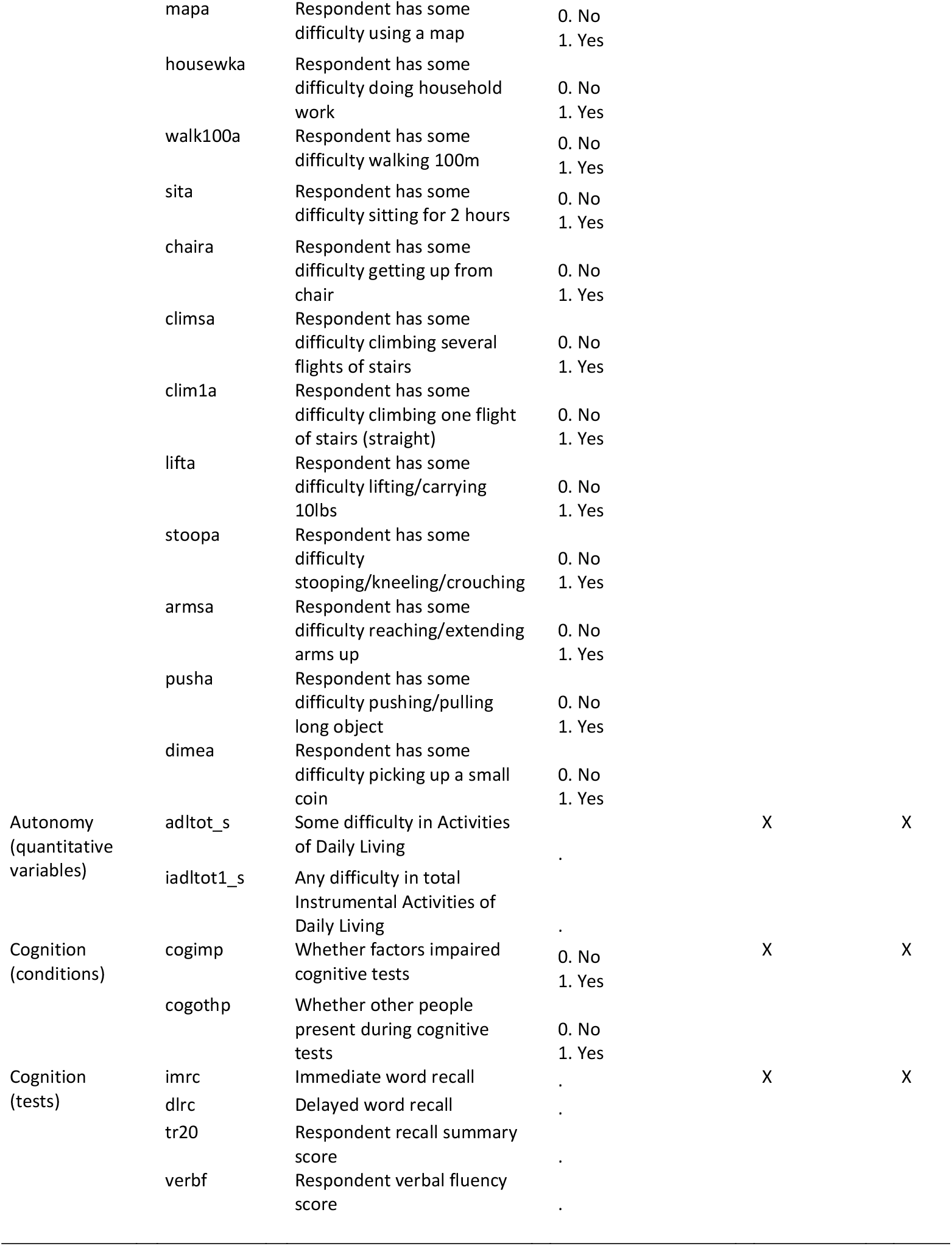
Summary of variables used for both imputation and Multiple Factor analysis (MFA) in ELSA.

**Supplementary Table 3:**
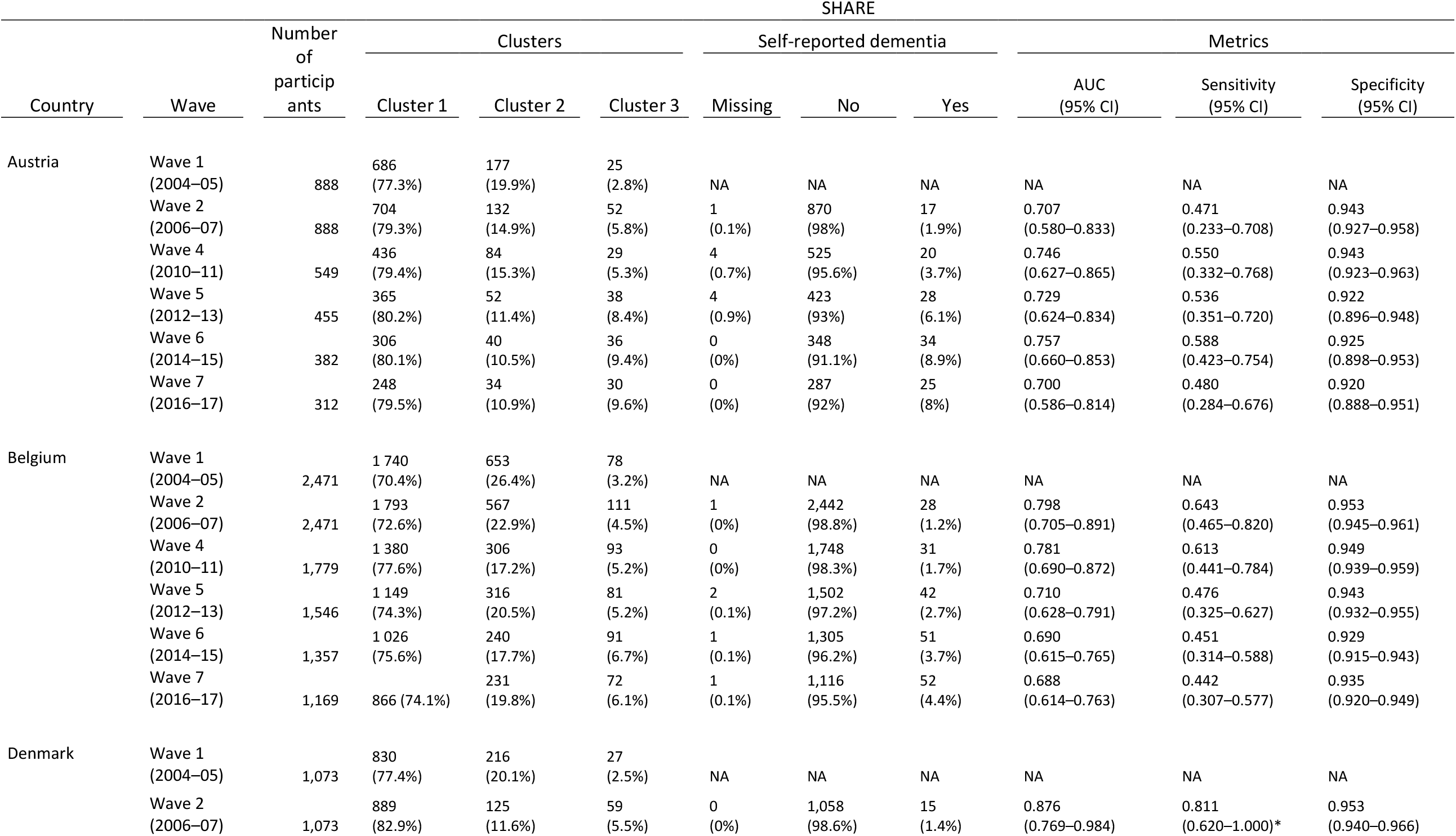
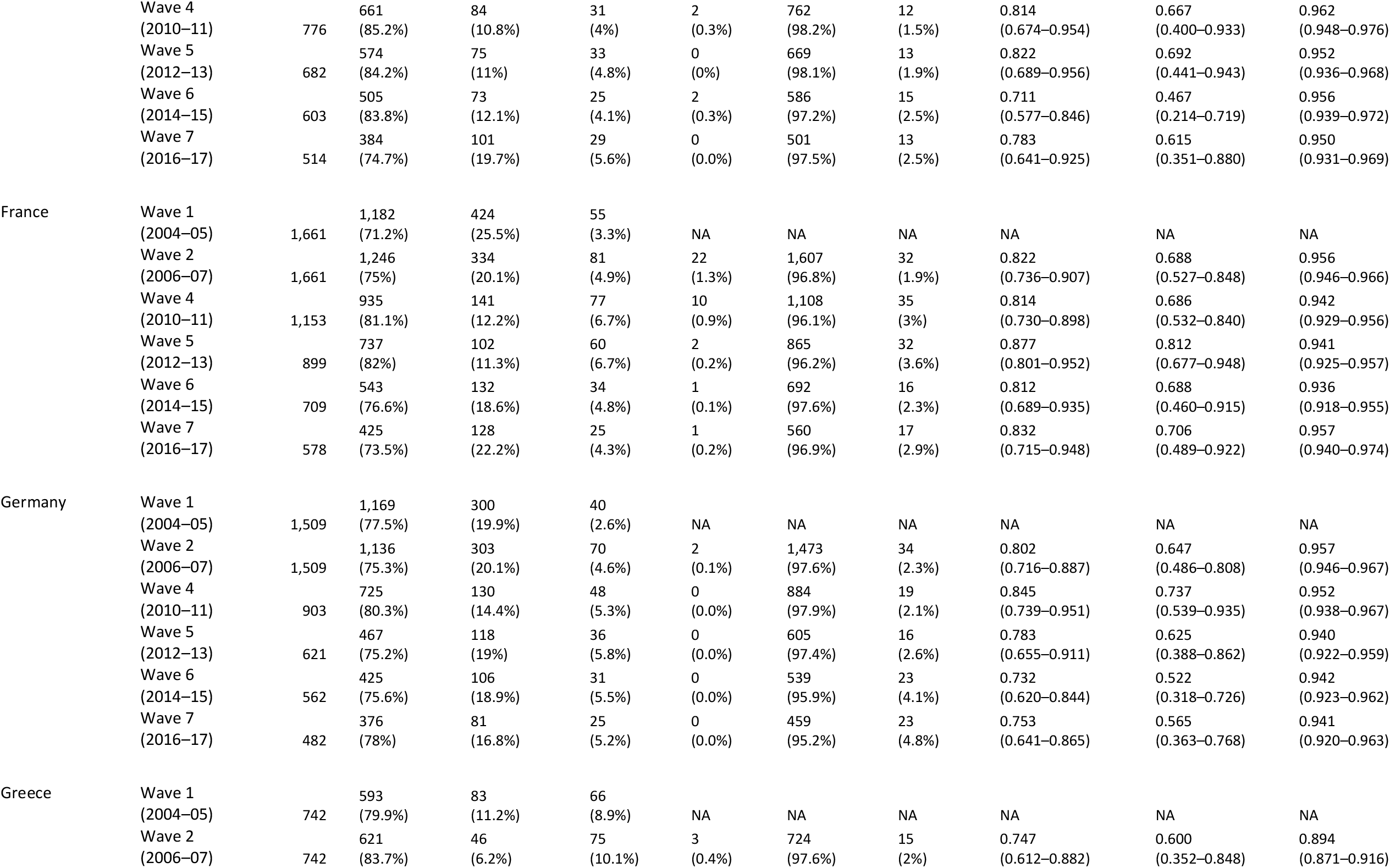
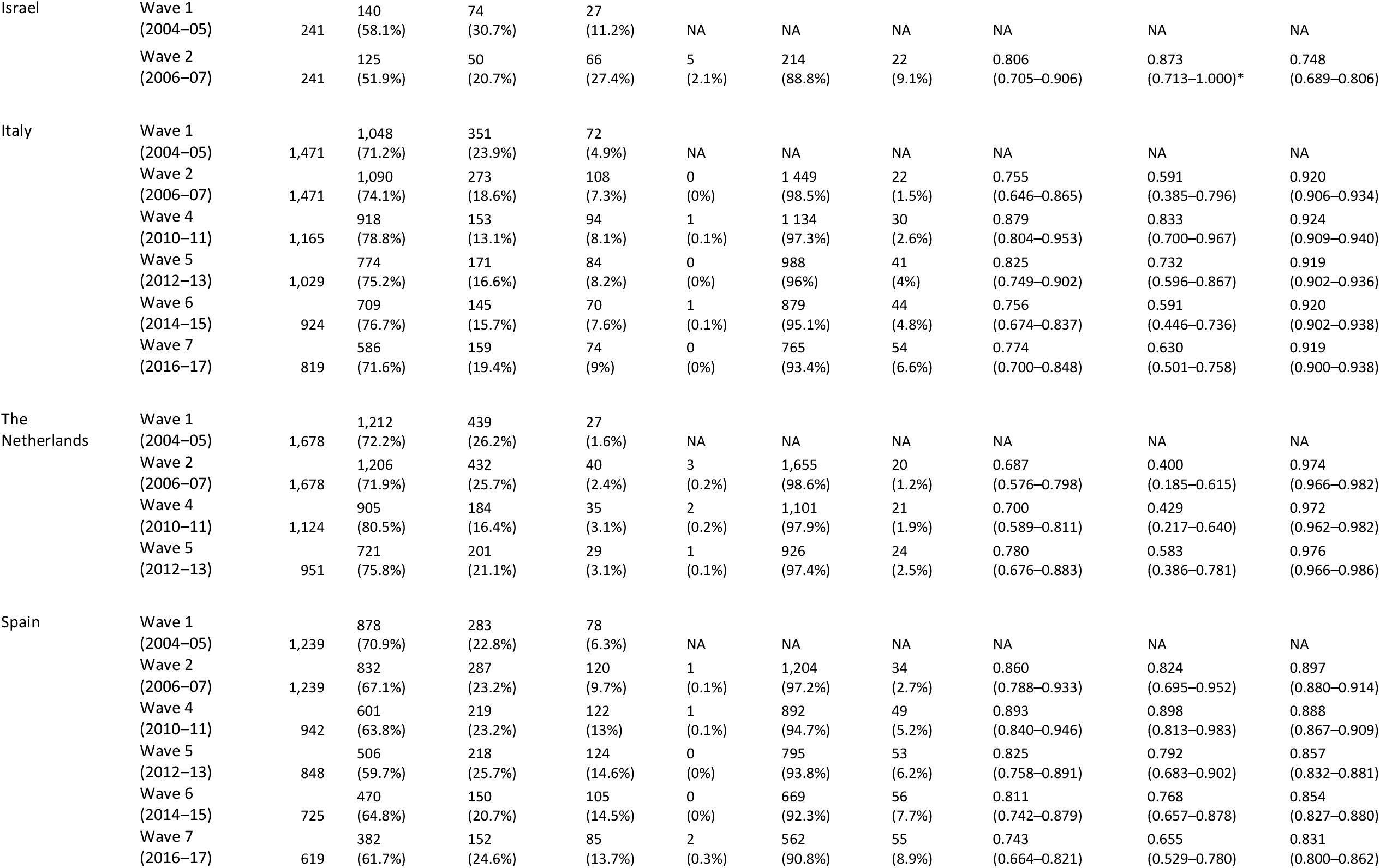
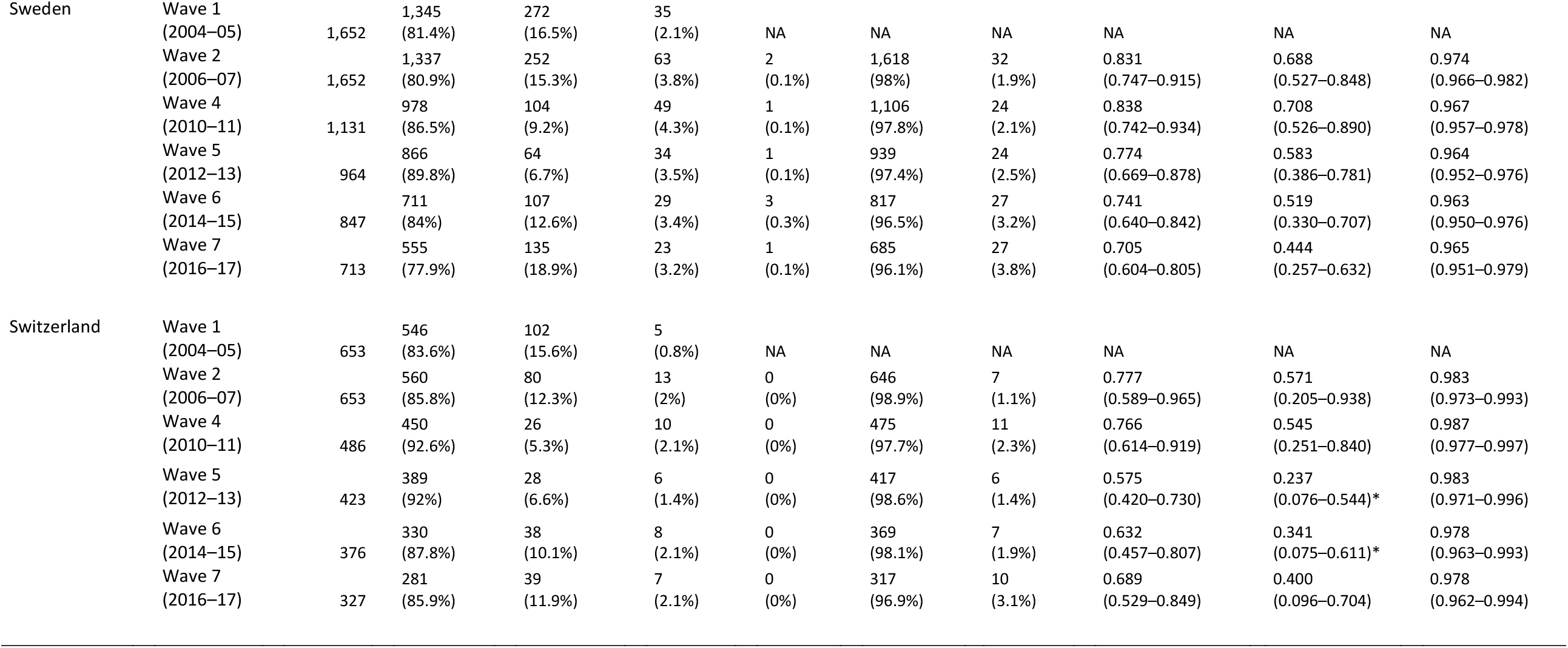
Self-reported dementia cases / Cluster 3 “Outcome” Comparison by country (SHARE), Abbreviations: NA, not available. *Values obtained using bootstrapping

**Supplementary Figure 2:**
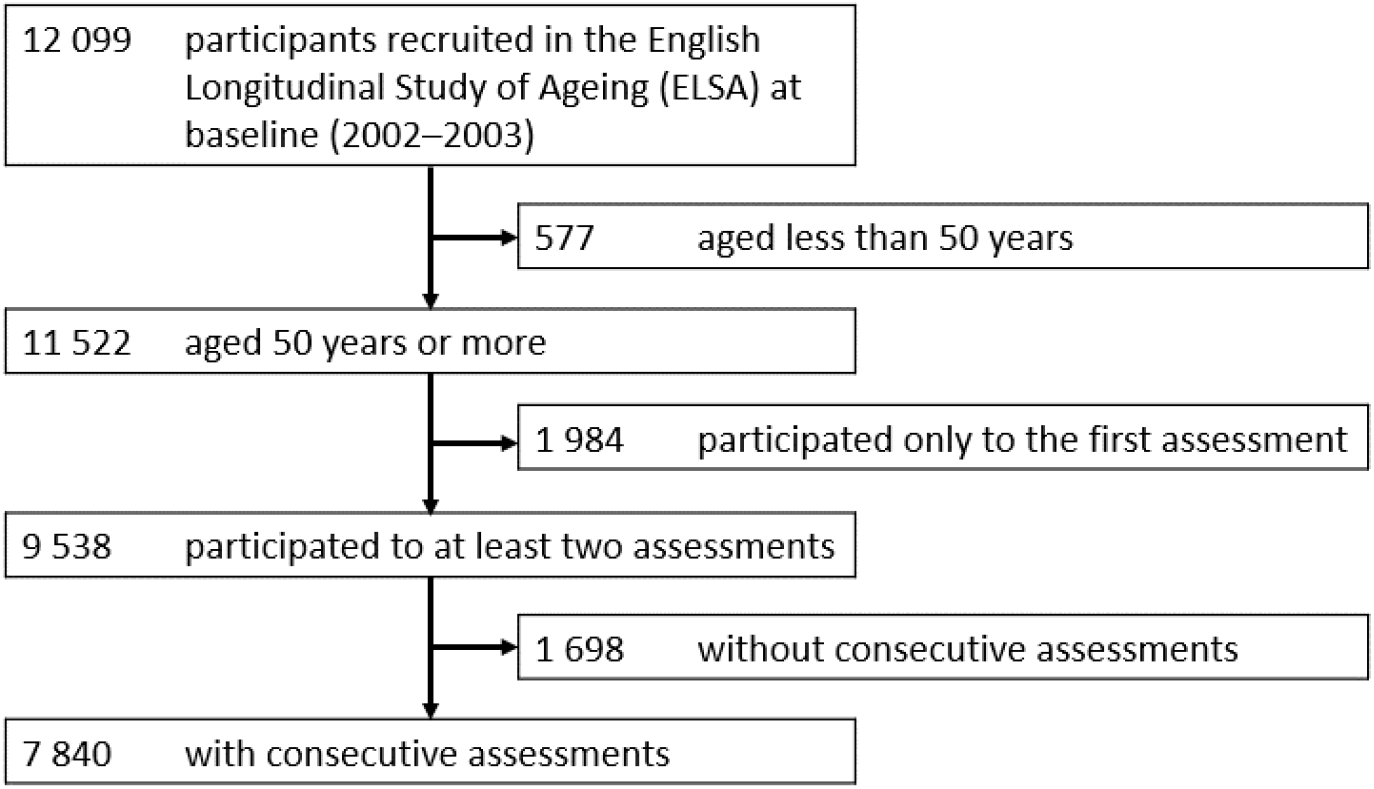
ELSA flow chart.

**Supplementary Table 4:** Baseline characteristics of the ELSA study participants according to the three clusters identified by the algorithm.

|  | ELSA |  |  |
| --- | --- | --- | --- |
|  | Cluster 1<br>(n=6,556) | Cluster 2<br>(n=625) | Cluster 3<br>(n=659) |
| Age, Years | 64.1 (9.7) | 65.8 (10.4) | 69.8 (11) |
| Sex |  |  |  |
| Female | 3,584 (54.6%) | 322 (51.5%) | 401 (60.8%) |
| Male | 2,976 (45.4%) | 303 (48.5%) | 258 (39.2%) |
| Education |  |  |  |
| Less than upper secondary education | 2,646 (40.4%) | 345 (55.2%) | 423 (64.2%) |
| Upper secondary and vocational training | 3,010 (45.9%) | 247 (39.5%) | 208 (31.6%) |
| Tertiary education | 900 (13.7%) | 33 (5.3%) | 28 (4.2%) |
| Mobility difficulty score [0–7] | 1.1 (1.4) | 1.6 (1.8) | 4.9 (1.4) |
| Autonomy |  |  |  |
| ADL score [0–6] | 0.1 (0.4) | 0.3 (0.7) | 2.7 (1.5) |
| IADL score [0–7] | 0.1 (0.4) | 0.3 (0.7) | 2.5 (1.4) |
| Cognition |  |  |  |
| Immediate Word Recall [0–10] | 5.7 (1.7) | 5.2 (1.8) | 4.6 (1.9) |
| Verbal Fluency [0–49] | 20.2 (6.1) | 18.7 (6.4) | 16.5 (6) |

**Supplementary Figure 3:**
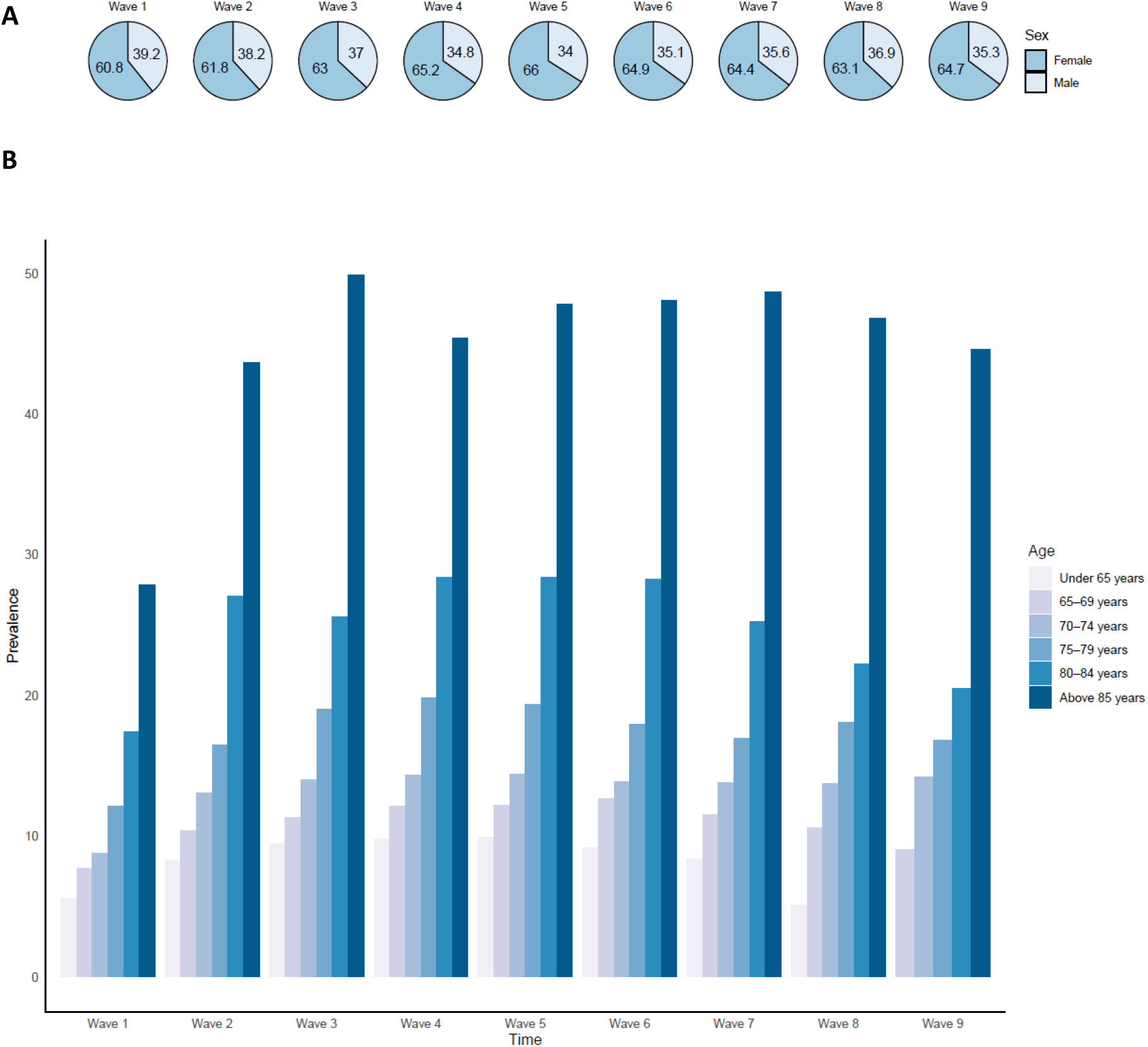
Prevalence of participants from the ‘‘Likely dementia’’ cluster by sex (3.A), and by age (3.B)

**Supplementary Table 5:** Comparison of self-reported dementia cases and Cluster 3 “Likely dementia” cases. Abbreviations: AUC, Area under the Curve; CI, confidence interval.

| ELSA |  |  |  |  |  |  |  |  |  |  |
| --- | --- | --- | --- | --- | --- | --- | --- | --- | --- | --- |
| Wave | Number of participants | Clusters |  |  | Self-reported dementia |  |  | Metrics |  |  |
|  |  | Cluster 1 | Cluster 2 | Cluster 3<br>(Likely Dementia) | Missing | No | Yes | AUC<br>(95% CI) | Sensitivity<br>(95% CI) | Specificity<br>(95% CI) |
| Wave 1<br>(2002–03) | 7,840 | 6,556<br>(83.6%) | 625<br>(8%) | 659<br>(8.4%) | 3<br>(0%) | 7,813<br>(99.7%) | 24<br>(0.3%) | 0.646<br>(0.546–0.746) | 0.375<br>(0.181–0.569) | 0.917<br>(0.911–0.923) |
| Wave 2<br>(2004–05) | 7,840 | 6,210<br>(79.2%) | 566<br>(7.2%) | 1 064<br>(13.6%) | 0<br>(0%) | 7,778<br>(99.2%) | 62<br>(0.8%) | 0.749<br>(0.685–0.813) | 0.629<br>(0.509–0.749) | 0.868<br>(0.861–0.876) |
| Wave 3<br>(2006–07) | 6,662 | 5,204<br>(78.6%) | 380<br>(5.7%) | 1,038<br>(15.7%) | 0<br>(0%) | 6,550<br>(98.9%) | 72<br>(1.1%) | 0.814<br>(0.762–0.866) | 0.778<br>(0.682–0.874) | 0.850<br>(0.841–0.859) |
| Wave 4<br>(2008–09) | 5,720 | 4,468<br>(78.1%) | 292<br>(5.1%) | 960<br>(16.8%) | 0<br>(0%) | 5,630<br>(98.4%) | 90<br>(1.6%) | 0.832<br>(0.788–0.877) | 0.822<br>(0.743–0.901) | 0.843<br>(0.833–0.852) |
| Wave 5<br>(2010–11) | 5,125 | 3,956<br>(77.2%) | 257<br>(5%) | 912<br>(17.8%) | 0<br>(0%) | 5,018<br>(97.9%) | 107<br>(2.1%) | 0.834<br>(0.793–0.874) | 0.832<br>(0.761–0.903) | 0.836<br>(0.826–0.846) |
| Wave 6<br>(2012–13) | 4,642 | 3,525<br>(75.9%) | 267<br>(5.8%) | 850<br>(18.3%) | 0<br>(0%) | 4,544<br>(97.9%) | 98<br>(2.1%) | 0.829<br>(0.786–0.872) | 0.827<br>(0.752–0.901) | 0.831<br>(0.820–0.842) |
| Wave 7<br>(2014–15) | 4,067 | 3,127<br>(76.9%) | 190<br>(4.7%) | 750<br>(18.4%) | 0<br>(0%) | 3,963<br>(97.4%) | 104<br>(2.6%) | 0.805<br>(0.759–0.851) | 0.779<br>(0.699–0.859) | 0.831<br>(0.820–0.843) |
| Wave 8<br>(2016–17) | 3,551 | 2,764<br>(77.8%) | 125<br>(3.5%) | 662<br>(18.7%) | 0<br>(0%) | 3,459<br>(97.4%) | 92<br>(2.6%) | 0.767<br>(0.714–0.820) | 0.707<br>(0.613–0.800) | 0.827<br>(0.815–0.840) |
| Wave 9<br>(2018–19) | 3,033 | 2,314<br>(76.3%) | 135<br>(4.4%) | 584<br>(19.3%) | 0<br>(0%) | 2,939<br>(96.9%) | 94<br>(3.1%) | 0.785<br>(0.734–0.836) | 0.745<br>(0.657–0.833) | 0.825<br>(0.811–0.839) |

**Supplementary Table 6:** Multistate models for the transition to cluster 3 (“Likely dementia”) Analyses using age as time-scale. All transitions were adjusted for sex. Transition towards the third cluster (“Likely dementia”) was further adjusted for age and each risk factor individually. All risk factors were taken at baseline. Main analysis was based on a multistate model (Model 1). In sensitivity analysis, cases identified either at the first or the second wave were removed (Model 2). Abbreviations: HR, hazard ratio; CI, confidence interval; NA, not available

| ELSA |  |  |  |  |
| --- | --- | --- | --- | --- |
|  | Main analysis |  | Sensitivity analysis |  |
|  | Model 1 (n=7,840) |  | Model 2 (n=6,784) |  |
|  | HR (95% CI)<br>(1 → 3) | HR (95% CI)<br>(2 → 3) | HR (95% CI)<br>(1 → 3) | HR (95% CI)<br>(2 → 3) |
| Education | **1.89 (1.64-2.17)** | 1.19 (0.77-1.82) | **1.6 (1.37-1.87)** | 1.07 (0.62-1.85) |
| Hearing | **1.75 (1.52-2.03)** | 1.35 (0.86-2.12) | **1.63 (1.39-1.91)** | 1.27 (0.7-2.29) |
| Hypertension | **1.42 (1.24-1.62)** | **1.64 (1.13-2.38)** | **1.39 (1.2-1.61)** | 1.3 (0.78-2.16) |
| Drinking (> 21 units) | **0.6 (0.43-0.83)** | 1.23 (0.68-2.24) | 0.79 (0.58-1.08) | 1 (0.37-2.7) |
| Smoking | **1.93 (1.6-2.33)** | 1.63 (0.98-2.71) | **1.18 (1.79-2.64)** | 1.7 (0.87-3.33) |
| Depression | **2.03 (1.74-2.37)** | **2 (1.26-3.17)** | **1.86 (1.57-2.19)** | 1.3 (0.69-2.42) |
| Social isolation | **1.61 (1.38-1.86)** | 1.52 (0.93-2.47) | **1.41 (1.2-1.65)** | 1.61 (0.85-3.06) |
| Physical inactivity | **2.65 (2.27-3.1)** | **2.69 (1.73-4.18)** | **1.74 (1.42-2.13)** | **2.02 (1.1-3.69)** |
| Diabetes | **1.77 (1.38-2.26)** | **2.23 (1.26-3.95)** | **1.62 (1.24-2.13)** | 1.84 (0.87-3.9) |
| Obesity | **1.53 (1.32-1.77)** | 0.97 (0.63-1.48) | **1.62 (1.38-1.9)** | 0.8 (0.44-1.46) |
| Pollution | NA | NA | NA | NA |

